# Bioelectrical Impedance Analysis in the nutritional assessment and prediction of complications in patients undergoing surgery for head and neck malignancies, A pilot observational study

**DOI:** 10.1101/2022.01.28.22269997

**Authors:** Lai Yi Ting, Peh Hui Yee, Hanis Binte Abdul Kadir, Lee Chun Fan, N. Gopalakrishna Iyer, Wong Ting Hway, Gerald Tay Ci An

**Affiliations:** Yong Loo Lin School of Medicine, Singapore; Dietetics, Singapore General Hospital, Singapore; Health Services Research unit, Singapore General Hospital, Singapore; Centre for Quantitative Medicine, Duke-NUS Medical School, Singapore; Department of Head and Neck Surgery, Singapore General Hospital, Singapore; Department of Head and Neck Surgery, National Cancer Centre, Singapore; Duke-NUS Medical School, Singapore; Health Services and Systems Research, Duke-NUS Medical School, Singapore

## Abstract

**Background:** Patients with head and neck malignancies are especially vulnerable to developing malnutrition, which has a significant impact on morbidity and mortality. Identification of high risk patients is hence critical for optimising outcomes.

**Objective:** It is hypothesised that bioimpedance analysis (BIA) can provide information on nutritional status and risk of perioperative complications in a timely and accurate manner. The study objectives are; to correlate BIA parameters with Subjective Global Assessment (SGA) scores, and determine the association of BIA parameters with common perioperative complications in patients undergoing head and neck surgery.

**Method:** This is a cohort study of 61 patients who were admitted for elective head and neck surgery from 2018-2019. Prior to surgery, patients were evaluated in a preoperative multidisciplinary allied health professional clinic for formal SGA scoring. Bioelectrical impedance analysis was performed using the Bodystat Quadscan 4000. One-way ANOVA and Fisher ‘s exact test were performed for associations between SGA and BIA parameters and receiver operating characteristic (ROC) curves were plotted for determination of optimal cut-off values of phase angle and Wellness marker in detecting malnutrition and perioperative pneumonia using Youden ‘s Index (YI).

**Results:** 45 males and 16 females with mean ± SD age of 62 ± 1.6 years old were included in the study. Significant differences were observed in Wellness Marker (p=0.004) and phase angle (p=0.006) amongst patients in the 3 SGA categories. BIA parameters (p=0.011 and p=0.032 for Wellness Marker and phase angle respectively) were associated with perioperative pneumonia. No significant differences were observed for other perioperative complications namely surgical site infections, salivary leak/fistula, and flap complications.

**Conclusion:** Bioelectrical Impedance Analysis is associated with Subjective Global Assessment and shows promise as a preoperative tool, in conjuction with SGA, to detect malnutrition in patients undergoing surgery for head and neck malignancies and highlight patients at risk of developing perioperative pneumonia.

## Introduction

Despite major advancements in healthcare, malnutrition remains ubiquitous in patients worldwide. Various studies have estimated worrying prevalence rates ranging from 20-50%.(1-3) In Singapore, up to one-third of admitted patients suffer from malnutrition.(4) The impact of malnutrition cannot be underplayed. Prolonged stays, increased readmission rates, treatment intolerance and higher in-patient and long-term mortality rates are known consequences in such patients globally.(5-7) Malnourished surgical patients have additional intraoperative and post-operative risks including: infections, delayed wound healing, impaired cardiorespiratory functions, and an increased risk of developing severe perioperative complications.(8) The culmination of these issues generate significant healthcare costs and reduce quality of life.(3)

Patients undergoing surgery for head and neck malignancies are notably vulnerable to developing malnutrition and up to 80% of patients with head and neck cancer are malnourished.(8) Common symptoms such as loss of appetite, xerostomia, taste alterations and dysphagia impair deglutition and mastication processes, leading to poor feeding and malnutrition.(9) Furthermore, inflammation and increased catabolism secondary to malignancy can also severely deplete muscle mass.(10)

Timely and accurate identification of patients at high risk for malnutrition is critical as it allows for early intervention for enhanced outcomes.(3) Therefore, there is a compelling need for robust, reliable and quick screening instruments to detect malnutrition. The Subjective Global Assessment (SGA), which relies on the nutritional history and clinical examination for a subjective impression of nutrition status has been a widely endorsed method of nutritional screening(11-13) and is considered by many to be the gold-standard. However, this method is subjective, time consuming and requires expertise.(14)

In recent years, bioimpedance analysis (BIA) has been lauded as a portable, safe, reproducible, inexpensive and non-invasive way of assessing body composition and nutritional status.(15) Specifically, BIA utilizes predictive equations via measurements of resistance (R) and reactance (Xc) of body tissues along with anthropometric data to generate the phase angle(16), a useful parameter reflective of body cell mass and cell membrane functions.(17) The Wellness marker, on the other hand, is a newly introduced parameter that calculates impedance ratio at various levels of frequency to reflect the generalized state of health of body cells without the need for predictive equations.

The favorable role of BIA in detection of malnutrition and prognosis have been demonstrated in various surgical patient populations such as in cardiac surgery(18), gastrointestinal surgery(19) and surgical cancer patients(20) internationally, however, no data exists on the use of bioimpedance for detecting malnutrition in patients undergoing surgery for head and neck malignancies in Singapore. Hence, the current study aims to determine the correlation between SGA scores and BIA parameters, as well as the correlation of BIA parameters with common perioperative complications in patients undergoing head and neck surgery.

## Materials and Methods

A prospective study was carried out on 61 patients out of a total of 97 patients scheduled for major head and neck oncological surgery in a tertiary hospital in Singapore from 2018 to 2019. These patients consented to the use of their personal data for research purpose and the study was approved by the institution ‘s ethics committee CIRB No. 2018/2234. Due to patient privacy policies of the institution, the authors are unable to give characteristics of those who refused to be included in the study.

Prior to surgery, patients were evaluated in a preoperative multidisciplinary allied health professional clinic. Subjective Global Assessment (SGA) scores were evaluated by certified dieticians on 61 patients prior to surgery and patients were identified as either well nourished, moderately malnourished or severely malnourished. Bioelectrical impedance analysis measuring phase angles and Wellness marker values was performed using the Bodystat Quadscan 4000 in 53 patients prior to surgery.

All patients were closely monitored and incidences of perioperative complications such as pneumonia, surgical site infections, salivary leak and/or fistula formation and flap complications were recorded until time of discharge.

Statistical analysis was performed with R (version 4.0.2). One-way ANOVA and Fisher ‘s exact test were used to test the association between the BIA parameters, SGA scores and development of perioperative complications such as pneumonia and surgical site infections. Patients who underwent flap surgeries and/or surgeries that had the potential for salivary leaks were analysed for the development of flap complications and salivary leak respectively. Receiver operating characteristic (ROC) curves were plotted to analyse the area under the curves (AUC). Optimal cut-off values of Wellness marker and phase angle for predicting malnutrition and perioperative pneumonia were obtained according to the Youden Index (YI).

## Results

### Patient characteristics

45 males and 16 females aged ranging from 25 to 88 years old were included in the study. The mean ± SD age was 62 ± 1.6 years old and the mean ± SD BMI was 23.3 ± 4.0.

Most patients (90.2%) were diagnosed with squamous cell carcinoma in the head and neck region. The remaining patients were diagnosed with adenoid cystic carcinoma, carcinoma ex pleomorphic adenoma, osteosarcoma, and undifferentiated nasopharyngeal carcinoma respectively.

### Prevalence of malnutrition

Based on the SGA scoring, 24 (39.3%) patients were found to be well-nourished, 32 (52.5%) patients were moderately malnourished, and 3 (4.9%) patients were severely malnourished.

Patient characteristics and nutritional status determined by SGA are summarised in **Table I**.

**Table I.** Baseline patient characteristics.

| Characteristics | N | Total | Mean ± SD<sup>1</sup> |
| --- | --- | --- | --- |
| Age | 61 | | 62 ± 1.6 |
| **Gender** | 61 |  |  |
| Male |  | 45 (73.8%) |  |
| Female |  | 16 (26.2%) |  |
| BMI | 61 | | 23.3 ± 4.0 |
| Length of stay | 61 | | 21 ± 19 |
| **Diagnosis** | 61 |  |  |
| Squamous cell carcinoma |  | 55 (90.2%) |  |
| Adenoid cystic carcinoma |  | 2 (3.3%) |  |
| Nasopharyngeal carcinoma |  | 2 (3.3%) |  |
| Carcinoma ex-pleomorphic |  | 1 (0.02%) |  |
| Osteosarcoma |  | 1 (0.02%) |  |
| **Neoplasm sites** | 61 |  |  |
| Oral cavity |  | 38 (62.3%) |  |
| Oropharynx |  | 7 (11.5%) |  |
| Nasal cavity |  | 3 (4.9%) |  |
| Nasopharynx |  | 1 (1.6%) |  |
| Larynx |  | 6 (9.9%) |  |
| Hypopharynx |  | 2 (3.3%) |  |
| Salivary gland |  | 2 (3.3%) |  |
| Thyroid |  | 1 (1.6%) |  |
| Middle ear |  | 1 (1.6%) |  |
| **T Staging** | 61 |  |  |
| T0 |  | 1 (1.6%) |  |
| T1 |  | 15 (24.6%) |  |
| T2 |  | 14 (23.0%) |  |
| T3 |  | 13 (21.3%) |  |
| T4 |  | 18 (29.5%) |  |
| **SGA<sup>2</sup> group** | 59 |  |  |
| Well nourished |  | 24 (40.6%) |  |
| Moderately malnourished |  | 32 (53.1%) |  |
| Severely malnourished |  | 3 (6.3%) |  |
| **BIA parameters** | 53 |  |  |
| Phase angle (°) | | | 5.95 ± 1.7 |
| Wellness marker (Hz) | | | 0.80 ± 0.05 |
| Fat mass (% of TBW3) | | | 29.0 ± 8.4 |
| Lean mass (% of TBW) | | | 71.0 ± 8.4 |
| Water mass (% of TBW) | | | 59.5 ± 6.7 |
| Body cell mass (kg) | | | 28.7 ± 6.4 |

## BIA parameters

The mean ± SD value of phase angle recorded was 5.96° ± 1.7 while the mean ± SD Wellness marker value was 0.80 ± 0.05. Other parameters recorded from BIA, including individual components of fat, muscle and water masses as a percentage of total body weight are summarised in Table I.

The mean ± SD duration of stay was 21 ± 19 days. In the course of their hospital stay, 21 (34.4%) patients developed perioperative complications. Specifically, 6 developed pneumonia, 9 patients developed surgical site infections, 2 patients developed salivary leak or had fistula formation and 7 patients had flap complications. None of the patients suffered acute myocardial infarctions or cerebrovascular accidents and there were no deaths recorded during admission.

Among the 3 SGA groups, there were statistically significant differences in phase angle (p=0.006) and the Wellness marker (p=0.004) measurements, as seen in **Table II**.

**Table II:** Phase angle and Wellness marker differences across SGA groups.

| Characteristic | Well-Nourished<br>N = 26<sup>1</sup> | Moderately malnourished<br>N=32<sup>1</sup> | Severely Malnourished<br>N=3<sup>1</sup> | p-value<sup>2</sup> |
| --- | --- | --- | --- | --- |
| Phase Angle (°) | 6.79 (1.53) | 5.41 (1.55) | 4.97 (1.32) | 0.006 |
| Wellness Marker (Hz) | 0.78 (0.05) | 0.82 (0.05) | 0.84 (0.04) | 0.004 |

<sup>1</sup> Mean (SD)
<sup>2</sup> One-way ANOVA

The optimal phase angle cut-off in predicting for moderate and severe malnutrition was below 4.7° (sensitivity 45.2%, specificity 95.5% YI: 0.406) while the optimal Wellness marker cut-off value in predicting for moderate and severe malnutrition was above 0.817. (sensitivity 60.0%, specificity 86.4% YI: 0.464). **Fig I** shows the receiver operating characteristic (ROC) curve of phase angle and Wellness marker for detecting malnutrition respectively. Both phase angle (AUC: 0.749) and Wellness marker (AUC: 0.755) provides fair diagnostic accuracy in identifying patients who are malnourished.

**Fig 1.**
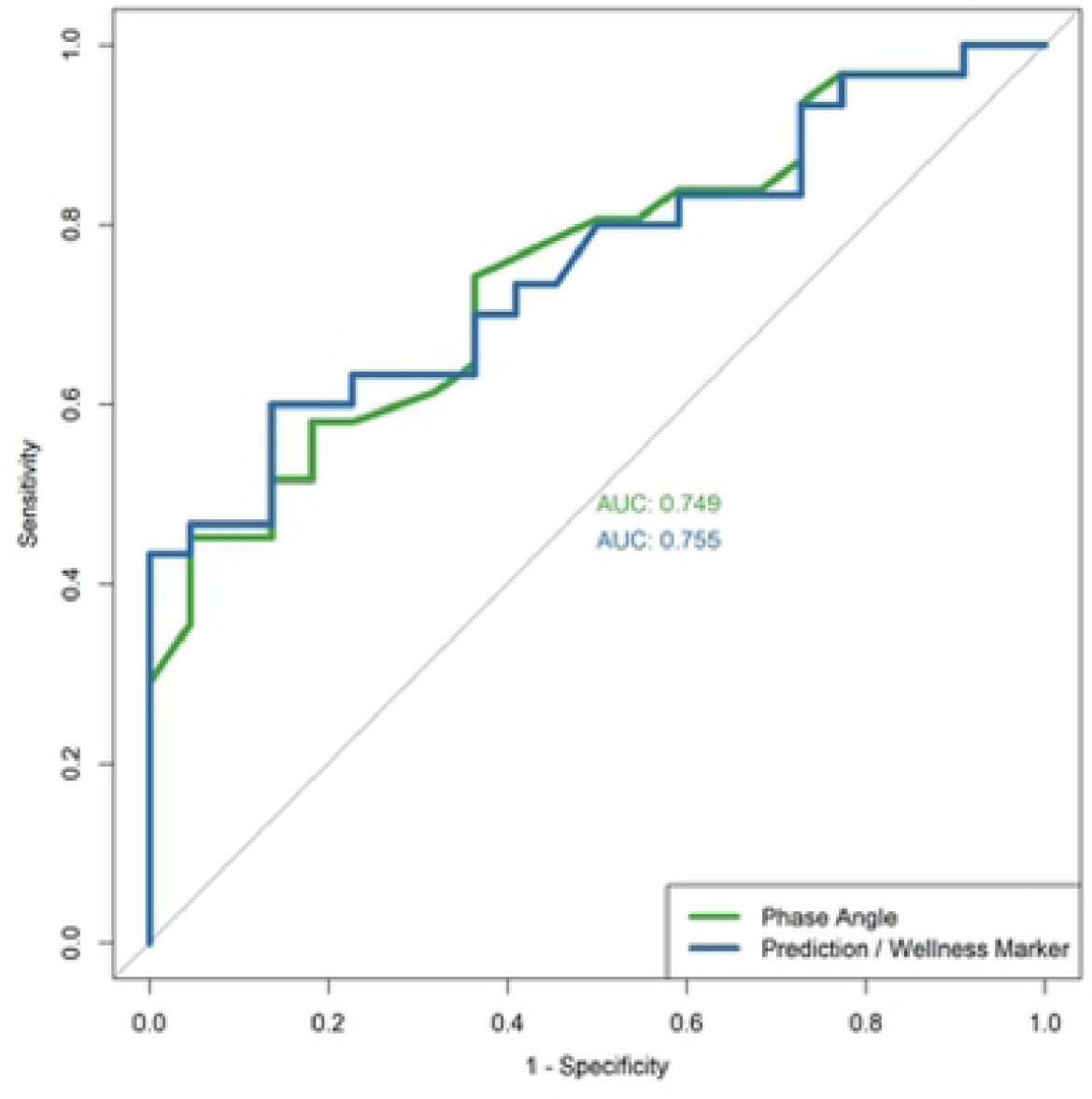
Receiver Operating Characteristic (ROC) curve of phase angle and wellness marker for detecting malnutrition. Both phase angle (AUC: 0.749) and Wellness marker (AUC: 0.755) provides fair diagnostic accuracy in identifying patients who are malnourished

Among all perioperative complications recorded, there were statistically significant differences in phase angle (p=0.032) and Wellness marker (p=0.011) values in patients who developed pneumonia. (**Table III**) No statistical differences in phase angle and Wellness marker was observed for the development of surgical site infections, salivary leak/fistula, and flap complications respectively.

**Table III:** Phase angle and Wellness marker differences in patients who developed perioperative complications.

| Perioperative complications | N | Yes<sup>1</sup> | No<sup>1</sup> | p-value<sup>2</sup> |
| --- | --- | --- | --- | --- |
| ***Pneumonia*** | 53 |  |  |  |
| Phase angle |  | 4.77 (1.25) | 6.26 (1.61) | 0.032 |
| Wellness marker (Hz) |  | 0.85 (0.02) | 0.79 (0.05) | 0.011 |
| ***Surgical Site Infections*** | 53 |  |  |  |
| Phase angle |  | 6.29 (0.91) | 6.06 (1.77) | 0.70 |
| Wellness marker (Hz) |  | 0.79 (0.03) | 0.80 (0.05) | 0.80 |
| ***Salivary Leak/Fistula*** | 34 |  |  |  |
| Phase angle |  | 5.95 (0.64) | 6.40 (1.57) | 0.70 |
| Wellness marker (Hz) |  | 0.80 (0.02) | 0.79 (0.05) | 0.70 |
| ***Flap Complications*** | 25 |  |  |  |
| Phase angle |  | 6.10 (1.10) | 6.26 (1.86) | 0.90 |
| Wellness marker (Hz) |  | 0.81 (0.04) | 0.79 (0.05) | 0.70 |

<sup>1</sup> Mean (SD)
<sup>2</sup> One-way ANOVA

The optimal phase angle cut off in predicting for perioperative pneumonia was below 5.5° (sensitivity 83.3%, specificity 69.8% YI: 0.531) while the optimal Wellness marker cut off was above 0.829 (sensitivity 100%, specificity 74.4% YI:0.744). **Fig II** shows the receiver operating characteristics (ROC) curve for phase angle and Wellness marker in predicting for development of perioperative pneumonia. Both phase angle (AUC 0.767) and Wellness marker (AUC 0.853) displayed good discriminating ability in predicting patients with perioperative pneumonia.

**Fig 2.**
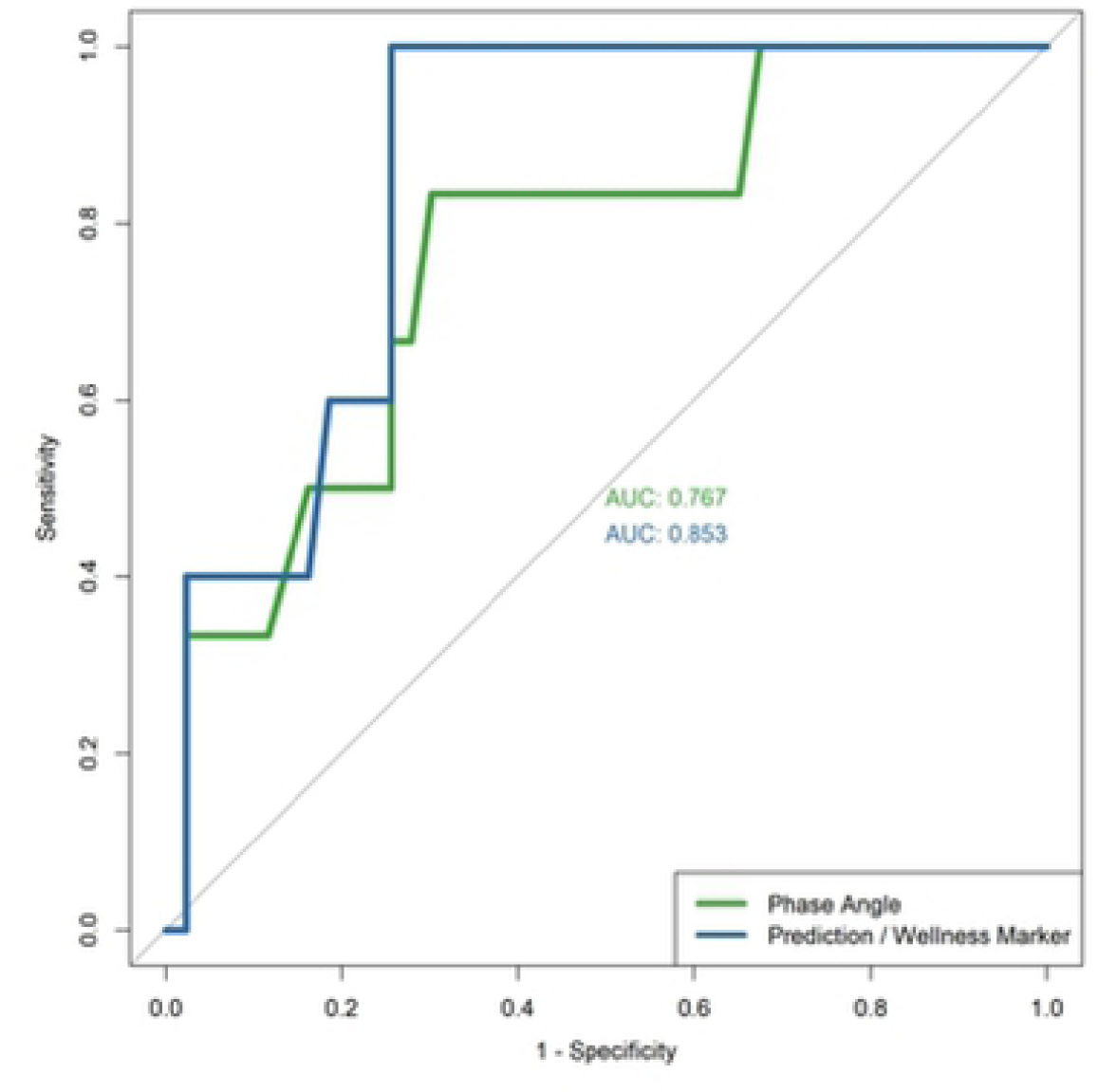
Receiver Operating Characteristic (ROC) curve of phase angle and wellness marker for predicting the development of perioperative pneumonia. Both phase angle (AUC 0.767) and Wellness marker (AUC 0.853) displayed good discriminating ability in predicting patients with perioperative pneumonia.

## Discussion

In our study, we found that a sizeable proportion of patients undergoing surgery for head and neck malignancies were malnourished (57.4%). Other studies have similarly reported high incidences of malnutrition in head and neck cancer patients, with prevalence rates ranging from 25% to 80%.(8, 21, 22)

Patients suffering from head and neck malignancies are especially susceptible to developing malnutrition because of impaired metabolism from disease processes, poor oral intake due to symptoms such as dysphagia, anorexia, mucositis, xerostomia and taste alterations.(23, 24) Furthermore, many patients suffer from chronic malnutrition associated with alcohol and tobacco use, compounding the present issue.(8)

Multiple cohort studies have demonstrated association between SGA scores and BIA parameters in healthy subjects(25), hospital in-patients(15) and surgical patients.(18, 20, 26) Notably, Małecka-Massalska *et al* demonstrated significantly lower phase angles amongst malnourished patients in a cohort of 75 newly diagnosed head and neck cancer patients.(27) Our results support these findings.

Bioimpedance analysis can provide valuable information on nutritional status via the determination of phase angles, which is a measure of cell membrane integrity and vitality and hence reliably reflects cellular health.(28) However, reference values of phase angles vary according to sex, age, BMI and disease processes.(29) Therefore, interpretation of phase angles should be population specific, as body composition varies between populations. For instance, higher body fat percentages in Asians compared to Caucasian counterparts of the same BMI presents significant challenges in comparing between studies. In the present study, we established optimal cut-offs specific to head and neck surgical patients in an Asian context. It may also be relevant for use in patients with head and neck squamous cell carcinoma, as these patients form the majority (90.2%) in our study population. Further studies will be required to validate and refine the values reported. The Wellness marker is a relatively newer parameter introduced only in more modern BIA systems and differs from phase angle measurements in that no predictive equations are required. Thus, Wellness marker values can be compared across populations as it is not affected by the subject ‘s weight, gender, age nor BMI values. To our knowledge, there are no other studies on malnutrition with Wellness marker measurements.

Our study also established that BIA predicts development of perioperative pneumonia. Several other studies have also demonstrated the role of BIA in predicting complications.(19) In our study population, BIA was predictive only for perioperative pneumonia amongst other complications studied, namely salivary leak, wound infections, and flap complications. BIA does not readily predict for the latter complications as they are often multifactorial and are likely more dependent on risk factors such as type of surgery, previous irradiation and other technical factors, which varies greatly between individuals. On the contrary, malnutrition has been strongly associated with pneumonia; protein calorie malnutrition has been found to impair pulmonary cell-mediated immunity processes and clearance of pathogens, resulting in increased incidence, severity and duration of pulmonary infections in malnourished individuals.(30) Besides perioperative complications, studies focused on head and neck cancer patients have also showed correlation of BIA parameters with prolonged hospital stay(31), survival rates(9, 10, 32), and radiotherapy outcomes.(33)

The value of BIA has evolved greatly since it was validated for use in assessing human body composition in 1983.(34) Since then, it has been well regarded as an objective, convenient, non-invasive, safe, portable and inexpensive tool for assessment of malnutrition and more.(35, 36) BIA may also be particularly useful in determining nutritional status and prognosis in cancer patients, as malignancy alter homeostatic processes and alters body composition.(37) However, BIA measures cannot be extrapolated to other populations and requires individuals to be relatively well hydrated for accurate measurements. (37) Due to limited validation studies in hospitalised patients and variability between BIA devices and body compartments estimated within studies, the prevailing American Society for Parenteral and Enteral Nutrition (ASPEN) clinical guidelines have not yet recommended BIA for use in clinical populations. (38)The SGA, on the other hand, is a validated method that is commonly employed in clinical settings. While it has achieved wide acceptance in its use globally(39), SGA lacks sensitivity to detect acute changes in nutritional status. (40, 41)Furthermore, it remains a subjective tool with inter-observer variability which greatly impairs its applicability on a continuum.(12, 42) Unlike BIA, SGA is also time consuming and requires trained professionals for reliable administration.(14) Therefore, we propose the use of BIA as a useful and convenient adjunct with other measures of nutrition in identifying and predicting for malnutrition and perioperative pneumonia.

The limitations of our study include a small patient cohort size from a single institution and the lack of long term follow up data. In addition, the type of peri-operative pneumonia and subsequent interventions these patients received have not been discussed within the scope of this paper. Further research may be required to determine cost effectiveness and practicality of BIA for routine use in an institutional setting.

## Conclusion

Bioelectrical Impedance Analysis is associated with Subjective Global Assessment, and can be used in assessing preoperative nutritional status for patients undergoing surgery for head and neck malignancies. BIA shows promise as a preoperative tool, in conjunction with SGA to detect malnutrition in patients undergoing head and neck surgery and highlight patients at risk of developing perioperative pneumonia.

## Data Availability

Data cannot be shared publicly because of patient privacy policies of the institution. Data are available from the SingHealth Centralised Institutional Review Board (CIRB) for researchers who meet the criteria for access to confidential data.

## Supporting Information

**S1 File. Strengthening the Reporting of Observational Studies in Epidemiology (STROBE) Checklist for cohort studies**.

